# Providing HIV Treatment during community-wide flooding: experiences of clients and Health Care Workers in Malawi

**DOI:** 10.1101/2024.04.28.24306432

**Authors:** M Mphande, R. Paneno, I Robson, K Phiri, M Cornell, JJ. van Oosterhout, J Njala, S Phiri, K Dovel

## Abstract

**Background:** In Malawi, community wide flooding, especially in high HIV burdened districts, continues to affect continuity of care and access to facilities. We explored the lived experiences of clients and healthcare workers (HCWs) to gain understanding of challenges and to propose interventions for improved ART care delivery.

**Methods:** Participants came six health facilities and surrounding communities impacted by flooding between Dec 2021-Apr 2022 in Chikwawa, Nsanje and Mulanje districts in Malawi. Facilities are supported by Partners in Hope, a local NGO and PEPFAR/USAID partner.

We conducted In-depth interviews with (IDIs) ART clients identified through medical chart reviews and focus group discussions (FGDs) with HCWs. IDIs and FGDs were coded using inductive and deductive methods in Atlas.ti.

**Results:** We conducted IDIs with 23 respondents, of which, ten were women, ten experienced treatment interruption (>28 days without medication) and 17 relocated from their homes. The Six FGDs involved 37 HCWs. (21 ART providers; 16 lay cadre).

In IDIs, most clients who relocated and lost livestock, possessions and ART medications. They travelling for income generation. Barriers to care included dangerous travel conditions, competing needs for time and fear of ill treatment at facilities. Some outreach clinics did not provide ART. Respondents were motivated to remain on care and motivators included fear of developing illnesses and HIV-status acceptance.

All providers said that lack of standardized guidelines affected preparedness and response and they advocated for guidelines, stakeholder coordination and adequate resources. Most also reported personal physical exhaustion, damage to their own houses and property, and drug stock-outs. Documentation due to loss of registers was also mostly mentioned.

**Discussion:** Clients motivated to remain in care but face barriers and challenges. National flooding protocols, adequate resource planning and seasonal 6-month ART dispensing may improve ART outcomes.

## Introduction

Geographical areas prone to extreme weather events also have the highest HIV prevalence.^1,2^ Further, extreme weather events, such as flooding disproportionately affect vulnerable populations, including those most at risk of HIV^1^. Extreme weather events cause migration and affect access to health services^3^. Like much of sub-Saharan Africa, Malawi is highly vulnerable to climate change due to poverty, being reliant on rain-fed agriculture, and having a limited flood-resistant infrastructure.^5–7^ Malawi is in the top five countries worldwide most affected by extreme weather events,^8^ and experienced three cyclones in the last 5 years - Idai in 2019, Anna in 2022 and Freddy in 2023 - that resulted in many human fatalities and had damaging effects by flooding on the environment and infrastructure. ^8,9^

Mitigating the impact of climate change associated events on health care delivery for people living with HIV (PLHIV) has therefore become critical in improving health outcomes. Destruction of infrastructure and transport hazards caused by severe flooding, interfere with accessing HIV care, resulting in treatment interruptions, thus increasing the risk of severe illness. ^2,10,11^ Within emergency camps, displaced PLHIV experience limited privacy, unwanted disclosure, stigma, and discrimination.^10^ However, some studies indicate that despite severe flooding, PLHIV are able to sustain ART adherence with adequate health system support. ^15,16^

Our programmatic data indicate that more clients interrupted treatment during flooding episodes in Malawi (Fig 1). However, studies that describe the provision of ART services during extreme weather events in Malawi are not available in the literature. Exploring client and health care worker (HCW) experiences, challenges and coping mechanisms during these events is crucial to designing interventions to prevent bottlenecks of service provision and increase access for clients and HCWs. In this paper we detail these experiences.

**Fig 1.**
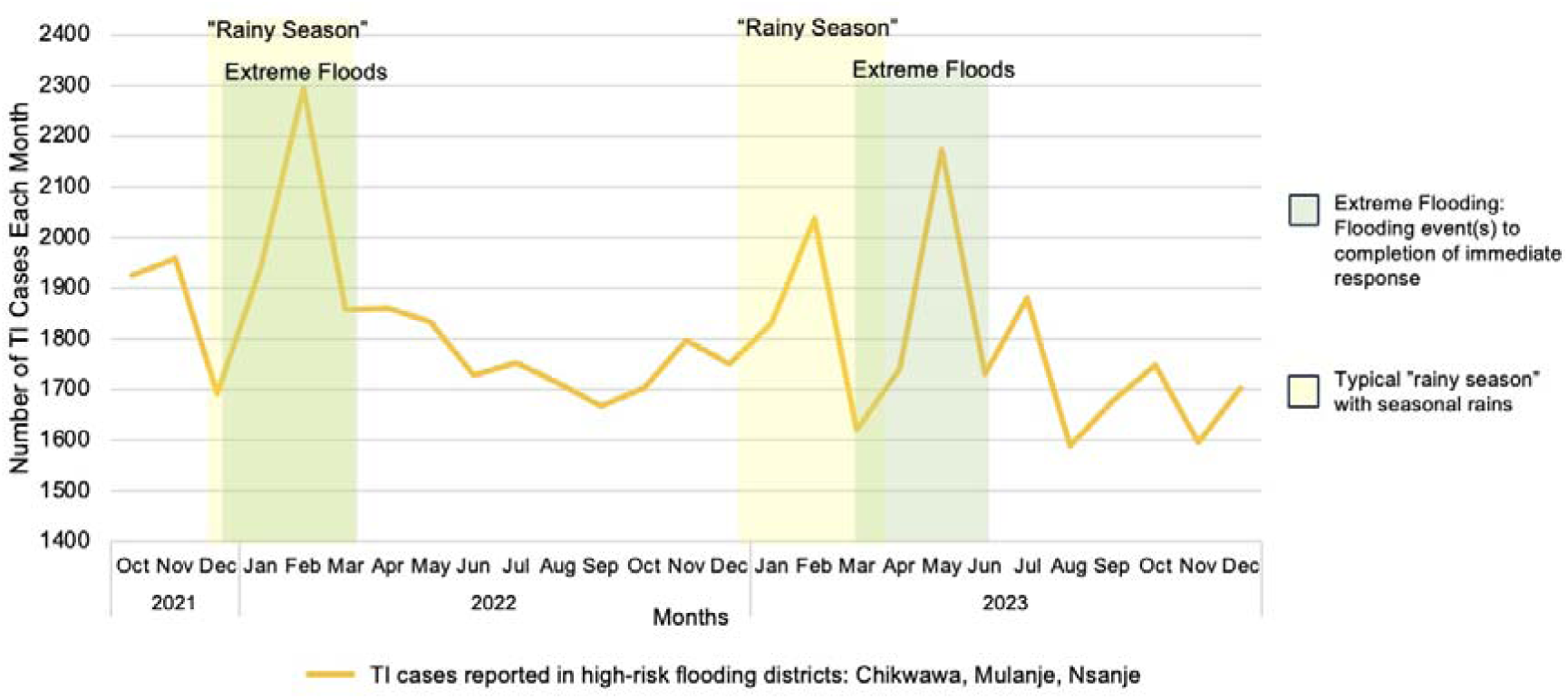
Treatment Interruption (TI) cases reported across Malawi Districts with highest flooding risk (three districts): Chikwawa, Mulanje and Nsanje Districts, 2021-2023

## Methods

We used qualitative methods to collect data on experiences during and immediately after flooding, with a particular focus on barriers to ART care, through semi-structured interviews with clients and focus group discussions with HCWs.

### Health facility and participant selection

We selected six health facilities that faced severe flooding between Dec 2021-Apr 2022: Lengwe and Kakoma health centres in Chikwakwa district, Chambe and Kambenje health centres in Mulanje district, and Mbenje and Ndamela health centres in Nsanje district. These facilities are supported by Partners in Hope (PIH), a Malawian faith-based non-governmental organization (NGO) and PEPFAR/USAID HIV care and treatment implementing partner.

We used routinely collected HIV program data within the selected facilities to identify facilities with a high number of ART clients that experienced treatment interruption (TI) during Dec 2021-Apr 2022. TI was defined as missing a scheduled ART clinic appointment for at least 28 days. Within these facility catchment areas, we selected 12 clients who had interrupted treatment during the period for in depth interviews. For each participant who interrupted treatment, we selected another a client residing in the same village who did not interrupt care for a total of 12 clients who did not interrupt treatment.

We conducted six focus group discussions with 16 providers (clinical officers and nurses) and 18 Treatment Supporters (TS; a lay cadre staff responsible for treatment continuation counseling and for tracing clients with TI) from the six facilities. We purposely identified Health Care Workers (HCWs) who had worked in the ART clinic at the identified sites during the flooding period.

### Data collection

#### In-depth interviews with clients

We conducted semi-structured in-depth interviews (IDIs), focused on lived experiences, health support received, and barriers and facilitators to ART care during flooding.

#### Focus group discussions with HCWs

We conducted focus group discussions (FGDs) with ART providers and TS to discuss HCW challenges, perceived health system challenges and solutions for ART retention and adherence during flooding.

#### Data analysis

We analyzed both IDIs and FGDs in Atlas.Ti 9 [4] using grounded theory [5]. We developed codes *a priori* based on existing literature about health care delivery and community health initiatives during flooding, and on literature about ART delivery in Malawi. We summarized codes and identified overarching themes in responses from both IDIs and FGDs.

## Results

### Demographics

Between July and August 2022, we conducted 23 IDIs with ART clients from six facility catchment areas impacted by flooding and conducted six FGDs with 34 HCWs who worked at these facilities (Table 1).

**Table 1.** Characteristics of ART clients who experienced flooding during 2021-2022 in southern Malawi.

| Characteristics | N=23 | % |
| --- | --- | --- |
| Sex |  |  |
| Female | 13 | 57% |
| Male | 10 | 43% |
| Median age, IQR | 40 | 28-46 |
| TI (*≥28 days late for ART appointment*) |  |  |
| Yes | 9 | 39% |
| No | 14 | 61% |
| Occupation of respondent |  |  |
| Farmer | 21 | 91% |
| Business | 2 | 9% |
| Relocated to camp |  |  |
| No | 6 | 26% |
| Yes | 17 | 74% |
| *Government camp* | 14 | 82% |

Among ART clients, 57% were female, the median age was 40 years and 39% experienced TI due to flooding. Due to the heavy flooding, 74% were displaced from their homes, with 82% of these individuals moving to government-supported camps.

Fifty percent of the HCWs were female and 53% were TS. All had worked during the flooding period. The average time they were employed at the flooded facility prior to the FGD was 27 months.

Flooding presented multiple challenges to ART engagement, both at the health systems- and the individual level. Clients and HCWs devised coping strategies to navigate these challenges, which we report below.

### General life disruption

#### Challenges and Coping Mechanisms

All ART clients (n=23) reported that life was disrupted due to flooding. Clients lost their homes, household belongings, and income generating activities due to flooding, including crops and livestock. Clients reported high levels of food insecurity.

> *“It was very hard for me to go to the facility and access my ART because I could not manage to cross the rivers. All seeds that I planted got washed away by floods, I also had goats and chickens which got washed away by floods as well.”- Female treatment interrupter*

Due to overarching life disruptions, many clients depended on NGOs and government donations, but these resources did not sufficiently meet basic housing, food and clothing needs. Some clients were forced to prioritize immediate and basic survival needs over accessing HIV treatment. Others argued that they could not take ART when they had food at home. Most clients had to travel to find work either in neighboring villages or across borders.

> *“Yes. I moved to Mozambique to earn a living. I could not come back to Malawi to get my treatment because I had no money for transport”- Male treatment interrupter*

More than half of HCWs also lost homes and were forced to relocate due to flooding. Almost all HCWs reported increased workloads and physical and psychological exhaustion during flooding. Providing routine health services at facilities and flood-specific services through outreach clinics was demanding and placed additional stress on HCW families who were also facing personal challenges. HCWs also reported not receiving aid from NGOs or from the government because they were not viewed as vulnerable to the floods but as part of the relief effort.

> *“We need support as health workers during this time, just like how the communities are supported. Like for instance, the previous floods, we were affected just like anyone and needed support to keep us going during this time.” – TS, Female*

#### Desired Solutions/Interventions

Given the desperate situation faced by clients and HCWs following flooding events, participants highlighted the need for a holistic response that addressed less visible issues such as the challenges of distant, rural residence, basic HCW needs, and mental health.

> *“An assessment should be done quickly to see the number of affected people and provide timely and necessary support. People from remote communities like us are abandoned. Support/aid for floods victims was focused on semi urban areas along the main roads leaving us with no hope and support” - female, treatment interrupter*

HCWs and clients believed that post-flooding economic recovery was needed, but rarely provided. Economic recovery programs would help flooding victims recover and enable people to move to highlands and reduce risk of repeated losses.

> *“…it would be helpful if well-wishers helped us with business capital so that we should be able to move from these flood-prone areas to highlands…If NGOs can’t help us, these problems will be there year after year because we don’t have anywhere to depend on” - male, non-treatment interrupter*

### Loss of ART medication during flooding

#### Challenges related to ART medication loss

Most clients (n=17, 77%) described losing all their medication either due to sudden flooding that washed away medication, or forgetting medication as they rapidly evacuated. More than two thirds of ART clients also lost personal medical records due to flooding, making it challenging to know ART appointment dates or to obtain the proper care when they were able to return to the clinic because facilities require the personal records to identify clients.

> *“Flooding affected my ART badly because they got washed away. My health passport got washed away. I couldn’t remember my appointment date, because I used to ask children to check my date from the health passport. I had no peace of mind and I didn’t know what to do so I just stayed”-female, treatment interrupter*

Flooding also caused frequent drug stock outs in facilities, with drugs being damaged at facilities or washed away, and facilities not receiving new drugs due to flooded roads. Given the increase in demand by clients who had lost their medication, providers had difficulty supplying medications both within facilities and at outreach clinics.

> *“Sometimes it might happen that the area affected by floods has a lot of ART clients and most client’s drugs have been washed away with floods, so that we used to have supply shortages. Because it happened that we took the drugs to the camps, we remained with a very few stocks at the facility, meaning other clients will get affected as well” -ART provider, male*

#### Desired Solutions/Interventions

Providers suggested that clients would benefit from standardized counseling curriculum on ART and health record storage so that they are easily accessible during flooding evacuation and kept dry. Providers also discussed the benefit of having increased stock in anticipation of seasonal flooding.

> *“Clients can be educated on what to do when their drugs get washed away or how to take care of their drugs and health passports during flooding*” -ART provider, female

### Limited physical access to health care

#### Challenge and Coping Mechanism

Both clients and HCWs reported that damaged roads, flooded rivers, and closed facilities made it difficult to access care and increased the risk of TI as clients were unable to collect refills.

> *“For me to miss my appointment it was because of the floods that cut so many rivers and the waters were running so fast and it was very hard for one to cross so we could just turn back and stay. This is what caused me to stay that long without going to the facility.”- female, treatment interrupter*

Facilities themselves were not spared from the impacts of flooding. Facilities within the flood zones were largely closed. In multiple facilities HCWs reported that the flooding caused loss of registers, thereby affecting care delivery. Identifying clients and their correct regimen, in the absence of electronic mobile records (due to power outage or unavailability of the electronic system) was difficult. One facility was filled with debris from flooding and clients needed to cross a flooded river to access the next closest facility.

> *“… we stayed for a very long time without electricity at the facility and most of the services needed electricity; for instance, TB samples needed electricity and it was hard to keep those samples and transportation to the DHO (District Health Office) could not be done right away. So, having these challenges, clients’ treatment was delayed.” -ART provider, male*

Even if a client was able to access the clinic, if ART pill bottles or client’s individual paper medical records (health passport) were lost, placing an individual on the correct regimen was a matter of guesswork.

> *“So let us say they lost their drugs and health* passports*, there are high chances of being given a different regimen, which is dangerous.” - ART provider, female*

Some clients were able to remain on treatment by traveling through dangerous conditions to get refills at camps and health facilities.

> *“Yes. I could swim across the river to get my ART. It was risky but there was nothing else that I could do to get my ART; so skillfully, I managed to swim across the river” - male, non-treatment interrupter*

Clients who were unable to undertake potentially dangerous travel and coped with loss of medications or inability to refill medications by borrowing from friends and relatives, although this was against guidelines.

> *“Yes. I started feeling unwell and I was risking my life for not taking ART for days so I found someone who lent me some ART…, She gave me pills that I used while figuring out means to get my ART at the clinic… I saw the colour and label of the bottle and the color of the pills”- female, non-treatment interrupter*

Some clients understood the dangers of sharing ART, and felt there was nothing they could do to resolve loss of ART until they could access care, resulting in TI.

At health system level, outreach clinics were set up in makeshift spaces within evacuation camps to address the growing need for ART care, in coordination with government and NGO support.

> *“It was hard for clients to move from the camps to the facility to get their treatment. So we had to introduce outreach clinics to help clients who missed their appointment and those who lost their drugs due to flooding” - ART provider, male*

#### Desired Solutions/Interventions

Both HCWs and clients recommended providing multi-month dispensation (MMD) prior to flooding-prone seasons to ensure clients have sufficient ART medication to mitigate the impact of flooding. Clients who lost their ART supply during flooding argued that 6-month MMD would address challenges related to mobility and lack of facility access. As the rains last for four months, they suggested having a 6-month supply would be crucial to adherence.

> *“It could also be better if clients were to be given enough ARTs before rainy season mostly December to March so that our clients should have to travel to health facilities as you are aware that most roads and rivers are impassable” - Treatment supporter-male*

Due to the decentralized medical record keeping, outreach clinics were not equipped to provide ART refills for clients with missing health records or those whose records were stored at another facility. HCWs suggested that a more centralized electronic medical record (EMR) may be valuable in synchronizing care between various sites for clients in disaster settings.

> *“With regards to EMR so clients’ information is accessed at any health facility if you just log in details so you will be able to cross-check with the client if s/he took ARVs from another facility.”-ART provider, male*

### Lack of privacy of ART services

#### Challenges and Coping Mechanisms

All clients and a few HCWs mentioned that the lack of privacy in overcrowded homes and camps during the flooding period presented major challenges for taking ART pills without being observed, and increased the chance of TI. Clients also faced a risk of unwanted disclosure when seeking care at outreach clinics within emergency camps.

> *“It becomes a challenge for clients to take ARVs even if they have some because of being afraid of unwanted disclosure. In camps, people stay together in tents that makes it difficult for clients to swallow ARVs as result they have days without taking ARVs due to lack of private space”-ART provider, female*

Despite this, some clients continued to take pills openly, accepting the possible unwanted disclosure and stigma.

> *” I did not feel shy. I could just take my bag, bring out ART and swallow without being worried of people. I did this because that is how I am and I could not hide it.” - female, non-treatment interrupter*

Providers needed to restructure services offered by outreach clinics and advertise carefully to prevent unwanted disclosure for clients.

> *” Seeing someone coming to where we set up the clinic they would conclude the person is on ART. So we had to tell them that we are there for different health services like HIV testing, so when a client comes in we also had to them that we also provide ART” – TS, female*

### Limited guidelines and health facility policies on flooding response

#### Barriers

Many HCWs noted the lack of national and local flood-preparedness guidelines and found existing ART and clinic operations guidelines inadequate for flood-related challenges. This led HCWs to rely on improvised strategies such as outreach clinics.

Providers struggled to meet patients’ needs while dividing resources between the facility and outreach clinics, due to limited staffing. They also encountered challenges in efficiently mobilizing limited Ministry of Health vehicles to adequately staff outreach clinics. Resupplying ART for clients according to standard protocols proved challenging due to reliance on phone calls with facilities to identify medication histories. This was hindered by high call volumes, paper records, and the lack of physical copies in outreach clinics, causing delays.

> *“If we refer from 2022 ART guideline there is no topic called ART in emergency. So it is important for providers to be oriented on this. Yes we do work to help clients but without following any guidelines we call it “phwanyaphwanya (willy-nilly)” - ART provider, male*

#### Solutions

Participants emphasized the importance of policies for clients on safeguarding and carrying their ART in case of evacuations. Additionally, participants felt clients would benefit from knowing that facilities had a non-blaming policy concerning lost ART or health records to reduce client reluctance to seek services. Participants underscored the necessity of addressing health record access in outreach clinics, proposing the implementation of electronic medical record (EMR) tools or transporting paper facility records to minimize reliance on confirmatory phone calls. Furthermore, they recommended incorporating contingency planning for low staffing and increased vehicle utilization during floods to optimize effectiveness of interventions.

> *“We have a very small number of providers so despite going for outreach clinics we also have to make sure to not compromise the services at the facility. So if there can be a possibility that during flooding there should be providers allocated special to work at the camps till flooding is over can be very helpful. We left very late from the outreach clinics because the vehicle would only drop us and leave to attend to other duties as well”. ART provider, male*

### Lack of a coordinated and comprehensive response

#### Challenges and Barriers

Given the lack of facility and governmental policies, HCWs provided ad-hoc services, juggling their clients’ needs with the resources available. Attempting to deliver services for flood victims, various departments, agencies and NGOs competed for shared facility resources such as vehicles and staffing. HCWs noted that within the camps, health service delivery frequently clashed with delivery of other services like food or clothing donations, which resulted in clients needing to choose between two desperately needed services.

> *“You could see WFP distributing food at the camps and at the same time you are there to provide ART services so most clients would prefer to go and receive food donations first then later on their ART. It was very costly because sometimes you could go and help very few clients because of that yet you have used a lot of fuel, time and the like.” - ART provider, male*

The lack of leadership on flooding response within facilities resulted in conflict between cadres of HCWs and disorganization in care delivery. For example, TS, who are responsible for tracing and client retention, felt excluded from community mobile clinic efforts when NGOs dictated the composition of the mobile clinic team, undermining their ability to trace clients.

> *“We need to have awareness where other NGOs would know or understand the roles and responsibilities of a TS, because they are not involved in outreach clinics yet we are the ones that are involved in counseling and tracing of clients. So a TS should always be involved in the whole process of organizing and implementing outreach clinics.” – TS, male*

#### Coordinated and Comprehensive Response Solutions

Providers advocated for advance planning with established leadership, and for clearly defined roles for all health facility staff during floods. They suggested that multiple services such as Under-5 clinic and primary care be delivered alongside ART services in outreach clinics, increasing resource utilization. They also suggested the need for better integration with NGO and governmental aid in emergency camps to avoid the need for clients to prioritize one urgent need over another.

> *“There should be a committee at facility level that should be put in place in order to respond. Through this committee there should be a person who will be responsible for organizing ART services at facility level and camps during floods” - treatment supporter, male*

## Discussion

In our study we identified in-depth insights into challenges, coping mechanisms, and proposed solutions to addressing TI amongst PLHIV in flood-prone regions of Malawi. Malawi is especially vulnerable to flooding given a heavy reliance on a predominantly rain-fed agricultural sector for its economy, employment, and food security^4^.

Our participants described extensive losses of crops, livestock, and property and many needed to relocate due damaged or destroyed residence. Loss of income-generating activity and resources resulted in significant challenges for ART clients, as household priorities (food, shelter, safety) conflicted with their engagement in care. HCWs in flooded regions were also heavily affected, losing their homes and property in floods. HCWs were not prioritized in relief efforts as they were seen as aid workers rather than those affected, impacting their ability to work effectively.

During evacuations from rising water, clients lost ART medication and health records, while facilities damaged by flooding also had losses of ART stores and records. Travel to health facilities became dangerous, impacting client ability to seek medication refills and government’s capacity to restock dwindling stocks. Outreach clinics were developed to address the rising need of ART in evacuation camps but lacked privacy and faced difficulty in providing the correct regimens due to impacted health record systems. Providers noted a lack of government and facility guidelines which would have helped to streamline and coordinate care delivery.

Due to disruptions of livelihoods, food insecurity becomes very important. Food security has been already been identified as a critical mechanism contributing to increased mobility of PLHIV and higher risk for other infections.^1,2^ Food insecurity is known to contribute to increased HIV transmission and poorer health outcomes for PLHIV due to worsening immune impairment, increasing an individual’s vulnerability to diarrheal disease.^2,3^. In this study, People Living with HIV (PLHIV) delineated an additional pathway through which flooding mediated food insecurity influenced HIV outcomes^19^. Clients expressed the belief that they should not consume ART on an empty stomach, fearing potential harm or adverse effects. This barrier to ART adherence in the absence of flooding has been extensively documented in literature from the region. PLHIV have consistently reported that ART intake exacerbates feelings of hunger, and share beliefs that taking medication without food can lead to significant medication effects.^4,5^ Consequently, individuals often face the dilemma of prioritizing immediate nutritional needs over adhering to their ART regimen. Providers noted the importance of addressing these frequently held beliefs and educating ART clients on the benefits of continuing ART even in the setting of food insecurity due to flooding. Overarching policies to integrate HIV care delivery with food aid to flooding survivors should be prioritized to reduce treatment interruption and health risks associated with flooding related food insecurity.

The relationship between clients and the health system agents that serve them in emergencies is often represented in a unidirectional, transactional way. HCWs are residents themselves of the flood prone regions and were vulnerable to the impacts of the floods. Health care workers are unable or unwilling to meet increasing clinical demands when they have unmet needs at home or experience increased risk of personal harm when going to work.^6^ Given the losses experienced by HCWs interviewed in our focus groups, as well as the dangerous travel required to come to work, staffing of clinics or outreach clinics was vulnerable to the individual experiences of these HCWs. Guaranteeing the safety and wellbeing of HCWs is critical to the success of any health system emergency response.^7,8^ The degree of health system resiliency (ability to prepare for and withstand) to public health emergencies is dependent on health workers^9^. Developing an aid plan that targets impacted HCWs during flooding will address some of the staffing problems and increase health system resiliency.

Medication and health record loss during flooding events is a significant challenge for flood survivors regardless of ultimate ability to remain in treatment^10^. Lost medications in the context of limited health system access resulted in coping strategies such as sharing medication or traveling through dangerous routes to access refills of medication. Globally, flood survivors have significant difficulty in accessing medications directly after climate emergencies because of local stock-outs and limited clinic access.^10^ Both clients and providers advocated for a dual strategy to tackle these obstacles: 1) Implementing Multi-Month Distribution (MMD) to supply six months’ worth of ART in advance of the rainy season’s flooding events. 2) Guaranteeing that facilities maintain sufficient and appropriately stored reserves of ART medications to pre-empt supply chain disruptions triggered by flooding.

Providing MMD has been shown to decrease burden on stretched health facilities by decreasing HCW workloads while maintaining viral load suppression, and was found to be useful in responding to emergencies such as the COVID-19 Pandemic.^11^ Improved coordination of local and international NGOs on both aid and medical supplies has been defined as crucial to the delivery of effective response in disaster settings^12–14^.

A lack of strategic guidelines and formalized body for leadership in planning and resource allocation impacted the ability for the HIV care delivery model to cope with the demands of the floods^14^. Providers shared that straightforward guidelines on the formation of an integrated flood response leadership group, involving stakeholders, would help to streamline aid and service delivery while reducing system redundancy^17^. One area where such coordination would be most impactful would be within the outreach clinic space. Outreach clinics were viewed as an acceptable alternative to access ART refills by some clients, but faced challenges in terms of provision of privacy and competing with clients who needed to be present and available to access other types of aid such as food aid or work opportunities^18^. Providers and clients noted that planning for a more private space for ART care delivery, as well as incorporation of other health services would decrease stigma. Coordinating ART outreach clinics with delivery of other aid would prevent clients from needing to prioritize one needed service over another.

### Limitations

Our study had methodological limitations. The qualitative approach does not enable us to capture distribution of experiences of clients and HCWs. However, our findings are crucial to building a model of important barriers, coping strategies, and potential solutions to ART delivery in flooding.

## Conclusion

In conclusion, considering climate related emergency in care provision is becoming critical to improving HIV treatment outcomes. Developing resilient healthcare systems relies upon supporting health care workers through systematic international and local support and coordination while recognizing local actors may themselves be impacted by the emergency. Its must be noted that developing a robust outreach clinic program is dependent upon coordination with other aid actors and provision of privacy for clients.

Our paper further argues that MMD, patient education, and pre-emptive stickups of medications may mitigate shortages and the impact of medication loss during floods. On a health system level. There is need for further research to understand client and HCW experiences can be incorporated into building a resilient health system.

## Acknowledgements

The authors would like to acknowledge Partners in Hope, its staff and clients from the PIH supported facilities for sharing their experiences and insights with the study team. The authors are also grateful to the research assistants, Mr Njala, Dr Cornell, Prof van Oosterhout and Prof Phiri for the guidance and for their support.

## Authors’ contributions

MM and KD conceptualized the study design. MM conducted qualitative data collection; MM supervised quantitative data collection. MM, RP and IR coded qualitative data; and MM, RP, KP, IR, and KD participated in qualitative data analysis. MM and PR drafted the initial manuscript. All authors reviewed the manuscript and supported with edits.

## Declarations

The study was supported under the ENGAGE trial, which is supported by the National Institute for Mental Health (R01-MH122308) and the National Institute of Health Fogarty International Center (K01-TW011484-01). KD is supported by Health Fogarty International Center. We are very grateful for the contributions of all participating HCWs and study participants

## Compliance with Ethical Standards

The study protocol is approved by the National Health Sciences Research Council (NHSRC) (#1099) in Malawi. There are no potential conflicts of interest to declare

## Disclaimer

The content is solely the responsibility of the authors and does not necessarily represent the official of Partners in Hope

## Data availability statement

The data that support the findings of this study are available from the corresponding author, MM, upon reasonable request.

Newer citations I need to loop in to the above that somehow didn’t sync up:

